# High prevalence of *Schistosoma mansoni* infection among adults with chronic non – communicable diseases in Malawi – a cross-sectional study at Mangochi District Hospital

**DOI:** 10.1101/2022.06.04.22275987

**Authors:** Wongani Nyangulu, Christina Sadimba, Joyce Nyirenda, George Twaibu, John Kamwendo, Kelvin Chawawa, Angella Masano, Elizabeth Chilinda, Sekeleghe Kayuni, Adamson S. Muula, Kenneth Maleta

**Affiliations:** Public Health and Nutrition Research Group, Department of Nutrition, Kamuzu University of Health Sciences, Blantyre, Malawi; Department of Tropical Disease Biology, Centre for Neglected Tropical Diseases, Liverpool School of Tropical Medicine, Liverpool, United Kingdom; MASM Medi Clinics Limited, Medical Aid Society of Malawi (MASM), Blantyre, Malawi; Department of Community & Environmental Health, School of Global and Public Health, Kamuzu University for Health Sciences, Blantyre, Malawi

**Author notes:** Corresponding author: Wongani Nyangulu.

**Keywords:** *Schistosoma mansoni*, Non-communicable diseases, Urine POC-CCA test, Kato-Katz microscopy

## Abstract

**Background:** Schistosomiasis is a parasitic infectious disease caused by flatworms of the *Schistosoma* genus. The global burden of schistosomiasis is high. In Malawi, schistosomiasis is among the top 20 causes of outpatient department visits in health facilities. Schistosomiasis is among the most important but neglected causes of non-communicable diseases (NCD) peculiar to tropical endemic settings. While much is known about the contribution of *S. haematobium* to the NCD burden in Malawi, the role of *S. mansoni* remains largely unknown.

**Methods:** We conducted a cross-sectional study at Mangochi District Hospital. Adults over 18 years diagnosed with NCDs (n = 414), admitted or attending weekly outpatient clinics were recruited between August 2021 and February 2022. Data were collected on sociodemographic characteristics, medical history, body weight, blood pressure, and fasting blood glucose. Stool and midstream urine were collected for Kato Katz (KK) microscopy and urine circulating cathodic antigen (CCA) tests respectively. We computed prevalence of *S. mansoni* as number of positive KK and CCA tests, each divided by total submitted samples. Univariate and multivariable logistic regression were done to evaluate risk factors of NCDs and association between *S. mansoni* infection and NCDs.

**Results:** We recruited 414 participants, mean age 57 years (SD 16), 67% of whom were female. Prevalence of *S*.*mansoni* based on urine CCA was 15% (95% CI 11 – 19) and 0% on KK microscopy. Hypertension was the most common condition with a prevalence of 85% (95% CI 81 – 89), followed by diabetes mellitus with a prevalence of 42% (95% CI 37 – 46) and heart disease with a prevalence of 3% (95% CI 2 – 5). *S. mansoni* infection was not significantly associated with hypertension (OR 1.2 (95% CI 0.5 – 3.1), p – value 0.47), diabetes (OR 0.6 (95% CI 0.3 – 1.1), p – value 0.117) or heart disease (OR 2.0 (95% CI 0.4 – 10), p – value 0.416).

**Conclusions:** We observed high prevalence of *S. mansoni* infection among adults in the study. This is within the range observed in children in Mangochi from 10 – 56.7%.

## Background

Schistosomiasis is a parasitic infectious disease caused by flatworms of the *Schistosoma* genus (1)(2). Humans are infected by larval forms of the parasite called cercariae in contaminated water sources (1)(2). Once infected, the disease manifests in three progressive stages namely cercarial dermatitis (swimmer’s itch), acute schistosomiasis (katayama fever) and chronic schistosomiasis (1)(3)(4). There are six schistosome species known to cause disease in humans (5). However, two species are prevalent in Malawi. *Schistosoma haematobium* causes urogenital schistosomiasis, and *Schistosoma mansoni* causes intestinal schistosomiasis (3)(6).

The global burden of schistosomiasis is high. Schistosomiasis transmission has been reported in 78 countries (5). Every year, over 220 million people are infected with the various schistosome species (7). There are over 280, 000 deaths annually due to schistosomiasis (8). It also causes disability on a massive scale accounting for over 29 million disability adjusted life years (DALYs) (8). The greatest burden of disease is observed in Africa and South America among poor communities with limited or no access to clean water sources for cooking, washing and bathing (9). Children under 14 years of age are also particularly vulnerable to infection (9).

In Malawi, schistosomiasis is among the top 20 causes of outpatient department visits in health facilities (10). Over 40 – 50% of the population is at risk of infection (10). *S. haematobium* is the most common parasite in Malawi with an average national prevalence of 50% (6). While less common, *S. mansoni* infection is increasing in prevalence. This shift is no less dramatic than along the shores of Lake Malawi where the prevalence has gone from 0% before 2014 to 34.3% (95% CI 27.9 – 41.3) among school children in Mangochi in 2017 (6)(11). Follow up studies done after this period establish the emergence of *S. mansoni* as an epidemic in Mangochi district with prevalence among school children ranging from 10 – 56.7% (12).

Schistosomiasis is among the most important but neglected causes of non-communicable diseases (NCD) peculiar to tropical endemic settings (13). In Malawi, the STEPS survey identified risk factors of NCDs including tobacco smoking (25% in men vs 3% in women), excessive alcohol intake (30% in men vs 4% in women), overweight (28% in women vs 16% in men), physical inactivity (13% in women vs 6% in men) and raised cholesterol (11% in women vs 6% in men) (14). However, these are common to both high and low – middle income countries. Chronic *S. haematobium* is associated with bladder cancer and cervical cancer, the 5^th^ and 2^nd^ most common cancers in Malawi (15)(16). Both *S. haematobium* and *S. mansoni* cause hepatosplenic disease with portal hypertension (2). Chronic *S. mansoni* infection also causes portal hypertension, pulmonary hypertension, cor pulmonale and eventual cardiac failure in 4 – 8% of those infected (17)(18).

While much is known about the contribution of *S. haematobium* to the NCD burden in Malawi, the role of *S. mansoni* remains largely unknown. Additionally, no local studies have been done to determine the prevalence of *S. mansoni* among adults who suffer from these NCDs. In this study, we investigated the role of *S. mansoni* infection in the NCD burden among adults with NCDs at Mangochi District Hospital. Our aim was to estimate the prevalence of *S. mansoni* infection in adults with NCDs at Mangochi District Hospital, evaluate the risk factors of NCDs in this population and determine if there was an association between *S. mansoni* infection and NCD syndromes.

## Methods

### Study design and population

We conducted a cross-sectional study enrolling adults over 18 years with an existing or recent diagnosis of NCD attending the NCD clinic or admitted at Mangochi district hospital. The study was done over a 7 month period from August 2021 to February 2022. The enrolment criteria were: 1) All consenting adults; 2) Age ≥ 18 years; 3) Existing or recent diagnosis of NCD. We excluded everyone with critical illness defined as in a comma or requiring mechanical ventilation.

### Study setting

The study was done in Mangochi, a lakeshore district at the southern tip of Lake Malawi. It has a population of 1, 148, 611 people, and 516, 976 are adults over 18 years old (19). Mangochi district hospital is the largest health facility in the district and serves as the major referral point for health centres and other facilities. It has an NCD clinic that operates twice a week on Tuesdays (diabetes clinic) and Thursdays (hypertension clinic). Recent studies in Mangochi district have demonstrated an emerging epidemic of *S. mansoni* infection amid the increasing prevalence of *S. haematobium* infections in this setting (12).

### Sample size and subject selection

To the best of our knowledge, there was no previous epidemiological data on prevalence of *S. mansoni* in adults with NCDs. Therefore, we used a conservative estimate of 50%. A sample size calculation with the single proportion formula (https://select-statistics.co.uk/calculators/sample-size-calculator-population-proportion/) showed that a sample of 376 was sufficient to estimate the prevalence of *S. mansoni* infection with 95% confidence and 5% margin of error. We estimated that there would be a refusal rate of 10%. Accounting for refusal, our final sample size was 414. We selected all participants who presented to the NCD clinic and the male or female wards at the hospital.

### Data and specimen collection

After collecting informed consent, research assistants administered a structured questionnaire and collected data on sociodemographic factors including education, marital and employment status, risk factors for non-communicable diseases including smoking and alcohol consumption, medical history, and results of relevant laboratory investigations. Then, participants were provided with a urine bottle and a stool collection bottle with a spoon to provide midstream urine and a stool sample. Those that could not provide samples on the same day provided them the following day.

### Urine circulating cathodic antigen (CCA) test

*S. mansoni* was detected in urine using the Schisto POC-CCA test (Rapid Medical Diagnostics, Pretoria, South Africa). Two drops of urine were transferred to the circular well of a test cassette delivering a total volume of 100µL. The result was read at 20 minutes. The presence of a control and test band was ‘positive’. The presence of a control band and the absence of the test band was ‘negative’. Participants who tested positive and reported back to the NCD clinic were prescribed praziquantel 40 mg/kg that they received at the district hospital pharmacy.

### Kato – Katz microscopy

To visualize Schistosoma ova in stool, fecal samples were pressed through a metal mesh (Sterlitech Corporation, Nylon screen, 100 mesh) to remove large particles. A portion of the sieved sample was then transferred to produce thick Kato-Katz smears on a slide. The smear was covered with a piece of cellophane soaked in a solution of glycerol and methylene blue. Light microscopy (x10 magnification and x40 magnification) was used to identify ova. Quantification of ova was not done.

### Statistical analysis

Data obtained from the study were entered into ODK and the output was in exported to Microsoft Excel spreadsheets. The spreadsheets were exported to STATA version 13 (StataCorp, 2013) and R version 4.1.1 (R Core Team (2021)) for analysis. We estimated the proportion of *S. mansoni* with binomial exact 95% confidence interval. Continuous variables were summarized using means ± standard deviations. Proportions were compared using two sample tests for comparing proportions and means were compared using the t-test. The association between two or more categorical variables was determined using the chi-squared (χ2) test. Univariate and multivariable logistic regression analysis were used to evaluate the risk factors of NCDs and the association between *S. mansoni* infection and NCD syndromes.

In the univariate logistic regression models, the outcome variables were binary: hypertension (yes/no), diabetes (yes/no) and heart disease (yes/no). The independent variables selected were *S. mansoni* infection, age, and sex, marital status, education status, and employment status, household income, smoking status, alcohol consumption and body weight. Risk factors with a p – value of < 0.05 on univariate analysis were selected for inclusion in the multivariable logistic regression model. The multivariable logistic regression models were checked using the Pearson χ2 goodness-of-fit test or the Hosmer – Lemeshow goodness of fit test as appropriate. If the p – value for the goodness of fit test was < 0.05, we rejected the model. If the p – value was > 0.05, we failed to reject the model and concluded that the model fitted well. We reported results with 95% confidence intervals and p – values with significance level set at p < 0.05.

### Ethical considerations

The protocol for the study was reviewed and approved by the College of Medicine Research Ethics Committee (COMREC P.10/20/3165). All participants provided written informed consent before participation in the study.

## Results

A total of 414 adults were enrolled into the study. The mean age was 57 years (SD 16) and 67% were female. There were statistically significantly more female than male participants recruited into the study with a difference in proportion of 34% (95% CI 24 – 44), p – value 0.000 (Table 1). The male and female participants had mostly similar age, educational background, marital status, employment status and average household income. However more men (78%) were currently married compared to women (51%) during the study period (difference of 27% (95% CI 16 – 38), p – value 0.0000). More men (45%) compared to women (29%) were also self-employed (difference of 16% (95% CI 0.1 – 32)). It was also noted that more women (49%) were unemployed compared to men (23%) during the study period (difference of 26% (95% CI 9 – 43), p – value 0.0085).

**Table 1.** Baseline sociodemographic characteristics of male and female participants

|  | Male | Female | Difference in mean/proportion (95% CI) | P – value |
| --- | --- | --- | --- | --- |
| N (%) | 137 (33%) | 277 (67%) | 34% (24 – 44) | 0.0000 |
| Age (years) |  |  |  |  |
| - Mean (SD) | 55 (18) | 58 (14) | 3 (-6 – 0.1) | 0.0538 |
| Education |  |  |  |  |
| - None | 21 (15%) | 67 (24%) | 9% (-27 – 9) | 0.3838 |
| - Less than primary | 58 (42%) | 140 (51%) | 9% (-24 – 6) | 0.2488 |
| - Primary school completed | 21 (15%) | 35 (13%) | 2% (-17 – 21) | 0.8344 |
| - Secondary school completed | 24 (18%) | 32 (12%) | 6% (-13 – 25) | 0.5288 |
| - College/University completed | 8 (6%) | 3 (1%) | 5% (-15 – 25) | 0.7254 |
| - Postgraduate degree | 5 (4%) | - | 4% | - |
| Marital status |  |  |  |  |
| - Never married | 11 (8%) | 7 (3%) | 5% (-15 – 25) | 0.6646 |
| - Currently married | 107 (78%) | 141 (51%) | 27% (16 - 38) | 0.0000 |
| - Separated | 3 (2%) | 20 (7%) | 5% (-4 – 18) | 0.7405 |
| - Divorced | 5 (4%) | 26 (9%) | 5% (-25 – 15) | 0.7089 |
| - Widowed | 11 (8%) | 83 (30%) | 22% (3 – 41) | 0.1243 |
| - Cohabiting | - | - |  |  |
| Work status |  |  |  |  |
| - Government employee | 11 (8%) | 12 (4%) | 4% (-15 – 23) | 0.6845 |
| - Non – government | 9 (7%) | 7 (3%) | 4% (-17 – 25) | 0.7219 |
| - Self employed | 61 (45%) | 81 (29%) | 16% (0.1 - 32) | 0.0491 |
| - Non paid worker | 8 (6%) | 21 (8%) | 2% (-22 – 18) | 0.8545 |
| - Student | 1 (1%) | - | - | - |
| - Home maker | 3 (2%) | 15 (5%) | 3% (-22 – 16) | 0.8190 |
| - Retired with benefits | 13 (9%) | 5 (2%) | 7% (-13 – 27) | 0.6034 |
| - Unemployed | 31 (23%) | 136 (49%) | 26% (9 – 43) | 0.0085 |
| Average household income (MK) |  |  |  |  |
| - Mean (SD) | 87 016.15 (84 493.59) | 69 117.52 (86 475.66) | 17 898.64 (-74 – 35 871) | 0.0509 |

A total of 339/414 (82%) participants submitted urine samples and 333/414 (80%) submitted stool samples. Prevalence of *S*.*mansoni* based on urine CCA was 15% (95% CI 11 – 19) and 0% on KK microscopy. A greater proportion of women (71%) submitted urine samples than men (29%), a statistically significant difference of 42% (95% CI 31 – 53), and p – value 0.0000. The prevalence of *S. mansoni* in men and women was 8% and 18% respectively. However, this difference was not statistically significant (10% (95% CI -32 – 12), p – value 0.4839). There were also no significant differences between participants who submitted stool and urine samples compared to those who did (Supplementary table 1).

Hypertension was the most common condition among the study participants with a prevalence of 85% (95% CI 81 – 89). This was followed by diabetes mellitus with a prevalence of 42% (95% CI 37 – 46). Some participants, 28% (95% CI 24 – 33) had both hypertension and diabetes mellitus. Heart disease with a prevalence of 3% (95% CI 2 – 5) was not significantly different between men (4% (95% CI 1 – 8)) and women (3% (95% CI 1 – 5)).

There was low reported prevalence of smoking in men (9%) and women (2%). There was also low reported prevalence of past alcohol consumption in men (9%) and women (1%). There was no statistically significant difference in smoking and alcohol consumption between the two groups (Supplementary table 2). Men and women had similar body weight (64 kg (SD 14) vs 64 kg (SD 18). However, height was not routinely measured to assess and compare mean body mass index. The mean systolic blood pressure in men was 150mmHg (SD 34) and 156mmHg (SD 33) in women (Table 2). The mean diastolic blood pressure in men was 84 mmHg (SD 17) and 88 mmHg (SD 17) in women. There was no statistically significant difference in blood pressure between men and women. Men had a mean fasting blood glucose (FBS) of 226 mg/dl (SD 114) and women had mean FBS of 250 mg/dl (SD 138). However, these were not statistically significantly different.

**Table 2.** Body weight, blood pressure and fasting blood glucose parameters of male and female participants

|  | Male | Female | Difference in means (95% CI) | P – value |
| --- | --- | --- | --- | --- |
| Body weight (kg) |  |  |  |  |
| - Mean (SD) | 64 (14) | 64 (18) | 0.1 (-4 – 3) | 0.9500 |
| Blood pressure (mm/Hg) |  |  |  |  |
| - Systolic Mean (SD) | 150 (34) | 156 (33) | 6 (-13 – 1) | 0.0983 |
| - Diastolic Mean (SD) | 84 (17) | 88 (17) | 4 (-7 – 0.02) | 0.0518 |
| Fasting blood sugar (mg/dl) |  |  |  |  |
| - Mean (SD) | 226 (114) | 250 (138) | 25 (-63 – 14) | 0.2057 |

In univariate and multivariable logistic regression analysis, the following variables were selected: *S*.*mansoni* infection, age, sex, education, marital status, employment status, average earnings, smoking, alcohol consumption and body weight. Age (OR 1.1 (95% CI 1.05 – 1.12), p – value 0.0000), completing primary school education (OR 0.2 (95% CI 0.05 – 0.98), p – value 0.047), non – government work (OR 7.2 (95% CI 1.2 – 42), p – value 0.028), being self-employed (OR 4.2 (95% CI 1.2 – 15), p – value 0.029), being unemployed (OR 4.7 (95% CI 1.2 – 18), p – value 0.025), and body weight (OR 1.04 (95% CI1.01 – 1.07), p – value 0.017) were significantly associated with hypertension on univariate and multivariable analysis (Supplementary table 3).

Age (OR 0.97 (95% CI 0.95 – 0.98), p – value 0.000), female sex (OR 0.4 (95% CI 0.2 – 0.7), p – value 0.001), smoking (OR 6.6 (95% CI 1.2 – 35), p – value 0.028), and body weight (OR 1.02 (95% 1.00 – 1.03), p – value 0.033) were significantly associated with diabetes on univariate and multivariable analysis (Supplementary table 4). The multivariable logistic regression models to evaluate risk factors of hypertension (Pearson χ2 goodness of fit p – value = 0.9942; Hosmer – Lemeshow χ2 p – value = 0.5344) and diabetes (Pearson χ^2^ goodness of fit p – value = 0.0563; Hosmer – Lemeshow χ^2^ p – value = 0.3820) were adequate and fit well with no evidence to reject the models. *S. mansoni* infection was not significantly associated with hypertension (OR 1.2 (95% CI 0.5 – 3.1), p – value 0.47), diabetes (OR 0.6 (95% CI 0.3 – 1.1), p – value 0.117) or heart disease (OR 2.0 (95% CI 0.4 – 10), p – value 0.416). Even after stratifying by sex to account for gender disparity in recruitment, *S. mansoni* infection was still not significantly associated with hypertension, diabetes or heart disease in men ((OR 0.5 (95% CI 0.1 – 2.4), p – value 0.412); (OR 0.7(95% CI 0.2 – 2.9), p – value 0.609) respectively) or in women ((OR 1.5(95% CI 0.4 – 5.2), p – value 0.556); (OR 0.7(95% CI 0.3 – 1.5), p – value 0.338); (OR 3.3(95% CI 0.5 – 20), p – value 0.205) respectively).

## Discussion

This is the first study in Malawi to estimate the prevalence of *S. mansoni* infection in adults with NCDs. There was high prevalence of *S. mansoni* infection in this population which is consistent with estimates in school going children in the district of 10 – 56.7% (12). This is also consistent reports of an epidemic of both *S. mansoni* and *S. haematobium* in the district.

We found no statistically significant association between *S. mansoni* infection and NCD syndromes of hypertension, heart disease and diabetes. However, previous studies have linked infection with development of pulmonary hypertension (20). Studies have also demonstrated the relationship between schistosomiasis and diabetes mellitus (21). Chronic *S. mansoni* infection causes vascular remodelling in the pulmonary circulation which leads to pulmonary hypertension and eventual heart failure. However, the pathological changes in the heart often lead to reduced cardiac output and therefore low systemic blood pressure or hypotension as opposed to hypertension. There are also recent findings that suggest parasitic infections could protect against autoimmune and inflammatory conditions such as diabetes mellitus (20)(22)(23)(24). In a Chinese study (n = 9539), adults over 40 years old with previous schistosome infection had lower prevalence of diabetes (14.9% vs 25.4%, p – value <0.001) (21). In adjusted logistic regression models, previous infection was protective of diabetes (OR 0.51 (95% CI 0.34 – 0.77) (21).

In this study, *S. mansoni* infection was detected using POC – CCA but not KK microscopy. While surprising, this is not an unusual occurrence. When prevalence of *S. mansoni* is very high (≥ 50%), results from POC – CCA and KK microscopy are almost equivalent (25). However, as prevalence declines below 50%, detection by POC – CCA becomes higher than KK microscopy by many degrees of magnitude (25). In previous studies, it was observed that when prevalence of *S. mansoni* using POC – CCA was less than 30%, prevalence using KK microscopy was below 10% and even 0% in some cases (25).

A limitation of our study was that the NCD syndromes were not defined a priori. We recorded diagnoses of hypertension, diabetes and heart disease made at the clinic and did not apply our own definitions which risks misclassification of these outcomes. Secondly, physical measurements which we captured and recorded in the study such as weight, systolic blood pressure, diastolic blood pressure and fasting blood glucose were not done by the study staff. We relied on measurements done at the clinic. To the best of our knowledge, these measurements are not done using standardized instruments which risks measurement bias. Another limitation was the lack of height measurements which were not done routinely at the clinic or on the wards. Therefore, we did not report body mass index (BMI) as a possible risk factor, using body weight as a proxy instead.

## Conclusions

There is a high prevalence of *S. mansoni* infection among adults with NCDs at Mangochi District Hospital. This reflects the high prevalence of infection observed in school going children in the district. However, there was no statistically significant association between *S. mansoni* infection and NCD syndromes. Due to the high prevalence of infection, we recommend continued screening and treatment of *S. mansoni* infection in this population. Larger follow up studies to evaluate the association between *S. mansoni* infection and NCDs including hypertension, pulmonary hypertension, heart disease and diabetes should be done to assess this major public health problem.

## Data Availability

All data produced in the present study are available upon reasonable request to the authors

## List of abbreviations

NCD: Non-communicable disease
ODK: Open Data Kit
POC-CCA: Point of Care-Circulating Cathodic Antigen
KK: Kato Katz
FBS: Fasting Blood Sugar

## Declarations

### Consent for publication

NA

### Availability of data and materials

The principal investigator will make data and materials available to interested parties on reasonable request.

### Authors’ contributions

WN designed the study. WN, CS, JN, GT collected data, stool and urine samples. EC and AM performed data entry. JK and KC performed laboratory tests on participant samples. WN performed statistical analysis. All authors read and approved the manuscript for publication.

## Acknowledgements

We are grateful to the staff and patients at Mangochi district hospital who participated in our study. We thank Asante Makuta and Alfred Kunje for the grants and logistical support during the conduct of this study.

The NCD BRITE consortium is supported by the National Heart, Lung, and Blood Institute of the National Institutes of Health under grant number 5U24HL136791. The content in this article is solely the responsibility of the authors and does not necessarily represent the official views of the National Institutes of Health. The authors report no relationships that could be construed as a conflict of interest.

