## Supplementary Table 1 for "High prevalence of *Schistosoma mansoni* infection among adults with chronic non – communicable diseases in Malawi – a cross-sectional study at Mangochi District Hospital"

Supplementary table 1. Comparison of participants who submitted and did not submit stool and urine samples

| | Submitted stool and urine specimens | | Did not submit stool and urine specimens | |
|---|---|---|---|---|
| Total sample size (414) | N = 339 | | N = 75 | |
| | Male | Female | Male | Female |
| N (%) | 137 (33%) | 277 (67%) | 38 (51%) | 37 (49%) |
| Comparison of socio-demographic characteristics | | | | |
| Age | | | | |
| - Mean (SD) | 55 (18) | 58 (14) | 58 (20) | 54 (13) |
| Education | | | | |
| - None | 21 (15%) | 67 (24%) | 8 (21%) | 9 (24%) |
| - Less than primary | 58 (42%) | 140 (51%) | 13 (34%) | 16 (43%) |
| - Primary school completed | 21 (15%) | 35 (13%) | 7 (18%) | 5 (14%) |
| - Secondary school completed | 24 (18%) | 32 (12%) | 7 (18%) | 6 (16%) |
| - College/university completed | 8 (6%) | 3 (1%) | 2 (5%) | 1 (3%) |
| - Post graduate degree | 5 (4%) | - | 1 (3%) | - |
| Marital status | | | | |
| - Never married | 11 (8%) | 7 (3%) | 4 (11%) | 2 (5%) |
| - Currently married | 107 (78%) | 141 (51%) | 27 (71%) | 20 (54%) |
| - Separated | 3 (2%) | 20 (7%) | 1 (3%) | 3 (8%) |
| - Divorced | 5 (4%) | 26 (9%) | 1 (3%) | 4 (11%) |
| - Widowed | 11 (8%) | 83 (30%) | 5 (13%) | 8 (22%) |
| - Cohabiting | - | - | - | - |
| Work status | | | | |

| | | | | |
|---|---|---|---|---|
| - Government employee | 11 (8%) | 12 (4%) | 2 (5%) | 2 (5%) |
| - Non – government employee | 9 (7%) | 7 (3%) | 4 (11%) | 1 (3%) |
| - Self employed | 61 (45%) | 81 (29%) | 16 (42%) | 11 (30%) |
| - Non – paid worker | 8 (6%) | 21 (8%) | 4 (11%) | 6 (16%) |
| - Student | 1 (1%) | - | 1 (3%) | - |
| - Home maker | 3 (2%) | 15 (5%) | - | 3 (8%) |
| - Retired with benefits | 13 (9%) | 5 (2%) | 4 (11%) | 3 (8%) |
| - Unemployed | 31 (23%) | 136 (49%) | 7 (18%) | 11 (30%) |
| Average household income | | | | |
| - Mean (SD) | 87 016.15 (84 493.59) | 69 117.52 (86 475.66) | 81 113.89 (67 342.91) | 63 654.05 (54 697.40) |
| Comparison of selected behavioral risk factors and medical history | | | | |
| Smoking | | | | |
| - Current smoker | 0 | 0 | 0 | 0 |
| - Ever smoked | 13 (9%) | 5 (2%) | 2 (5%) | 0 |
| Alcohol consumption | | | | |
| - Ever consumed | 12 (9%) | 4 (1%) | 3 (8%) | 0 |
| - In the past 12 months | 8 (6%) | 1 (0.4%) | 3 (8%) | 0 |
| - Stopped for health reasons | 8 (6%) | 2 (0.7%) | 1 (3%) | 0 |
| Cardiovascular disease | | | | |
| - Previous heart attack, chest pain (angina) or stroke | 8 (6%) | 13 (5%) | 2 (5%) | 2 (5%) |
| - Currently taking aspirin to prevent or treat disease | 33 (24%) | 82 (30%) | 7 (18%) | 9 (25%) |

| | | | | |
|---|---|---|---|---|
| - Currently taking regular statins to prevent or treat | 24 (18%) | 54 (20%) | 7 (18%) | 6 (16%) |
| Comparison of biological risk factors and anthropometry | | | | |
| Body weight (kg)<br>- Mean (SD) | 64 (14) | 64 (18) | 61 (14) | 64 (23) |
| Blood pressure (mm/Hg)<br>- Systolic Mean (SD)<br>- Diastolic Mean (SD) | 150 (34)<br>84 (17) | 156 (33)<br>88 (17) | 151 (39)<br>82 (17) | 150 (32)<br>85 (15) |
| Fasting blood glucose (mg/dl)<br>- Mean (SD) | 226 (114) | 250 (138) | 213 (105) | 292 (157) |
| Comparison of prevalence of NCD syndromes | | | | |
| Hypertension, % (95% CI) | 77% (69 – 83) | 90% (85 – 93) | 82% (66 – 92) | 84% (68 – 94) |
| Diabetes, % (95% CI) | 59% (50 – 67) | 33% (27 – 39) | 61% (43 – 76) | 35% (20 – 53) |
| Heart disease, % (95% CI) | 4% (1 – 8) | 3% (1 – 5) | 5% (0.7 – 18) | 5% (0.7 – 18) |
