## Supplementary Table 2 for "High prevalence of *Schistosoma mansoni* infection among adults with chronic non – communicable diseases in Malawi – a cross-sectional study at Mangochi District Hospital"

Supplementary table 2. Behavioural risk factors and medical history of male and female participants

| | Male | Female | Difference in proportions (95% CI) | P - value |
|---|---|---|---|---|
| Smoking | | | | |
| - Current smoker | 0 | 0 | | |
| - Ever smoked | 13 (9%) | 5 (2%) | 7% (- 13 – 27) | 0.6034 |
| Alcohol consumption | | | | |
| - Ever consumed | 12 (9%) | 4 (1%) | 8% (-11 – 27) | 0.5871 |
| - In the past 12 months | 8 (6%) | 1 (0.4%) | 5.6% (-149 – 26) | 0.8149 |
| - Stopped for health reasons | 8 (6%) | 2 (0.7%) | 5.3% (-148 – 25) | 0.7570 |
| Raised blood pressure | | | | |
| - Told by doctor in the past 12 months | 115 (84%) | 260 (94%) | 10% (3 – 17) | 0.0019 |
| - Taken medications for treatment in past 2 weeks | 20 (15%) | 36 (13%) | 2% (-17 – 21) | 0.8349 |
| Diabetes | | | | |
| - Told by doctor in the past 12 months | 84 (62%) | 91 (33%) | 29% (15 – 43) | 0.0001 |
| - Taken medications for treatment in past 2 weeks | 74 (54%) | 88 (32%) | 22% (7 – 37) | 0.0047 |
| Raised cholesterol | | | | |
| - Told by doctor in the past 12 months | 5 (4%) | 17 (6%) | 2% (-23 – 19) | 0.8636 |
| - Taken medications for treatment in past 2 weeks | 2 (1%) | 3 (1%) | 0 (-10 – 12) | 1.0000 |
| Cardiovascular disease | | | | |
| - Previous heart attack, chest pain (angina) or stroke (CVA) | 8 (6%) | 13 (5%) | 1% (-19 – 21) | 0.9214 |
| - Currently taking aspirin to prevent or treat heart disease | 33 (24%) | 82 (30%) | 6% (-24 – 12) | 0.5181 |
| - Currently taking regular statins to prevent or treat heart disease | 24 (18%) | 54 (20%) | 2% (-21 – 17) | 0.8366 |
| Chronic kidney disease | | | | |
| - Ever been told by doctor | 1 (0.7%) | 4 (1%) | 0.3% | - |
| - Ever been on dialysis or had kidney transplant | 0 | 1 (0.3%) | 0.3% | - |
