## Supplementary Table 3 for "High prevalence of *Schistosoma mansoni* infection among adults with chronic non – communicable diseases in Malawi – a cross-sectional study at Mangochi District Hospital"

Supplementary table 3. Regression analysis evaluating risk factors of hypertension

| Variable | Crude OR (95% CI) | P – value | Adjusted OR (95% CI) | P – value |
|---|---|---|---|---|
| S. mansoni | | | | |
| - Positive | 1.2 (0.5 – 3.1) | 0.47 | - | - |
| Age | 1.1 (1.05 – 1.09) | 0.000 | 1.1 (1.05 – 1.12) | 0.000* |
| Sex | | | | |
| - Male | 1 | | | |
| - Female | 2.6 (1.5 – 4.5) | 0.001 | 2.1 (0.96 – 4.6) | 0.063 |
| Education | | | | |
| - None | 1 | - | - | - |
| - Less than primary | 0.2 (0.1 – 0.8) | 0.024 | 0.7 (0.2 – 2.6) | 0.552 |
| - Primary school completed | 0.1 (0.03 – 0.5) | 0.004 | 0.2 (0.05 – 0.98) | 0.047* |
| - Secondary school completed | 0.1 (0.02 – 0.3) | 0.000 | 0.4 (0.1 – 2.0) | 0.284 |
| - College/University completed | 0.1 (0.01 – 0.3) | 0.001 | 0.8 (0.1 – 7.5) | 0.819 |
| - Postgraduate degree | 0.1 (0.01 – 1.7) | 0.121 | - | - |
| Marital status | | | | |
| - Never married | | - | - | - |
| - Currently married | 1 | 0.007 | 0.3 (0.1 – 1.4) | 0.120 |
| - Separated | 3.9 (1.5 – 10.5) | 0.015 | 0.5 (0.1 – 5.1) | 0.571 |
| - Divorced | 8.4 (1.5 – 47) | 0.036 | 0.4 (0.1 – 3.2) | 0.401 |
| - Widowed | 3.1 (1.1 – 15.8) | 0.000 | 0.4 (0.04 – 3.4) | 0.382 |

| | | | | |
|---|---|---|---|---|
| | 18 (4.6 – 70.1) | | | |
| Work status | | | | |
| - Government employee | - | - | - | - |
| - Non – government | 4.0 (0.9 – 18) | 0.071 | 7.2 (1.2 – 42) | 0.028* |
| - Self employed | 4.7 (1.9 – 12) | 0.001 | 4.2 (1.2 – 15) | 0.029* |
| - Non paid worker | 7.9 (1.9 – 34) | 0.005 | 2.9 (0.4 – 21) | 0.297 |
| - Student | - | | | |
| - Home maker | - | | | |
| - Retired with benefits | - | | | |
| - Unemployed | 6.7 (2.6 – 17) | 0.000 | 4.7 (1.2 – 18) | 0.025* |
| Average earnings | 0.9 (0.9 – 0.9) | 0.032 | 1.0 (1.0 – 1.0) | 0.117 |
| Smoking | | | | |
| - Never smoked | 4 (1.5 – 10.8) | 0.006 | 0.5 (0.1 – 3.4) | 0.476 |
| Alcohol | | | | |
| - Never used | 4.9 (1.8 – 13.7) | 0.002 | 4.7 (0.7 – 32) | 0.117 |
| Body weight | 1.18 (1.0 – 1.04) | 0.093 | 1.04 (1.01 – 1.07) | 0.017* |

NB: Pearson $\chi2$ goodness of fit p – value = 0.9942; Hosmer – Lemeshow $\chi2$ p – value = 0.5344, (*) statistically significant
