## Supplementary Table 4 for "High prevalence of *Schistosoma mansoni* infection among adults with chronic non – communicable diseases in Malawi – a cross-sectional study at Mangochi District Hospital"

Supplementary table 4. Regression analysis evaluating risk factors of diabetes mellitus

| Variable | Crude OR (95% CI) | P – value | Adjusted OR (95% CI) | P – value |
|---|---|---|---|---|
| S. mansoni | | | | |
| - Positive | 0.6 (0.3 – 1.1) | 0.117 | - | - |
| Age | 0.96 (0.95 – 0.97) | 0.000 | 0.97 (0.95 – 0.98) | 0.000* |
| Sex | | | | |
| - Male | 1 | | | |
| - Female | 0.3 (0.2 – 0.5) | 0.000 | 0.4 (0.2 – 0.7) | 0.001* |
| Education | | | | |
| - None | 1 | - | - | - |
| - Less than primary | 1.6 (0.9 – 2.7) | 0.105 | 1.2 (0.7 – 2.3) | 0.500 |
| - Primary school completed | 1.9 (0.9 – 3.8) | 0.076 | 1.3 (0.6 – 3.0) | 0.484 |
| - Secondary school completed | 4.2 (2.1 – 8.6) | 0.000 | 2.1 (0.8 – 5.4) | 0.117 |
| - College/University completed | 11 (2.3 – 56) | 0.003 | 3.5 (0.5 – 25) | 0.215 |
| - Postgraduate degree | 3.7 (0.6 – 24) | 0.159 | 1.3 (0.1 – 19) | 0.867 |
| Marital status | | | | |
| - Never married | 1 | - | - | - |
| - Currently married | 0.3 (0.1 – 0.8) | 0.019 | 0.4 (0.1 – 2.3) | 0.345 |
| - Separated | 0.2 (0.04 – 0.6) | 0.009 | 0.5 (0.1 – 3.0) | 0.423 |
| - Divorced | 0.2 (0.1 – 0.8) | 0.019 | 0.5 (0.1 – 2.9) | 0.424 |
| - Widowed | 0.1 (0.02 – 0.3) | 0.000 | 0.3 (0.1 – 1.8) | 0.195 |
| Work status | | | | |
| - Government employee | 1 | - | - | - |

| | | | | |
|---|---|---|---|---|
| - Non – government | 0.8 (0.2 – 3) | 0.773 | 0.7 (0.1 – 3.0) | 0.595 |
| - Self employed | 0.6 (0.2 – 1.5) | 0.278 | 0.97 (0.3 – 3.0) | 0.954 |
| - Non paid worker | 0.4 (0.1 – 1.2) | 0.103 | 0.6 (0.1 – 1.1) | 0.509 |
| - Student | - | | | |
| - Home maker | 0.2 (0.05 – 0.7) | 0.017 | 0.5 (0.1 – 2.9) | 0.471 |
| - Retired with benefits | 0.6 (5.4 – 2.2) | 0.487 | 0.1 (0.2 – 3.7) | 0.797 |
| - Unemployed | 0.3 (0.1 – 0.8) | 0.012 | 0.02 (0.2 – 2.2) | 0.523 |
| Average earnings | 1.0 (1 – 1.0) | 0.034 | 1.0 (1.0 – 1.0) | 0.953 |
| Smoking | | | | |
| - Never smoked | 1.9 (0.7 – 5.4) | 0.242 | 6.6 (1.2 – 35) | 0.028* |
| Alcohol | | | | |
| - Never used | 0.4 (0.1 – 1.1) | 0.089 | 0.6 (0.1 – 2.9) | 0.528 |
| Body weight | 1.01 (1.0 – 1.03) | 0.016 | 1.02 (1.00 – 1.03) | 0.033* |

NB: Pearson $\chi^2$ goodness of fit p – value = 0.0563; Hosmer – Lemeshow $\chi^2$ p – value = 0.3820;
(*) statistically significant
